# How do social norms influence the sexual and reproductive health of very young adolescents in sub-Saharan Africa? A scoping review

**DOI:** 10.1101/2025.08.05.25333028

**Authors:** Fardawsa A Ahmed, Owen Nyamwanza, Alice Ladur, Jermaine M Dambi, Frances M Cowan, Webster Mavhu

## Abstract

In sub-Saharan Africa (SSA), very young adolescents (VYAs, aged 10–14 years) have the worst sexual and reproductive health (SRH) outcomes of this age group worldwide due to behavioural, structural, socioeconomic and sociocultural factors, including social and gender norms. SRH programming often focuses on older adolescents (aged 15–19 years), overlooking VYAs. This scoping review sought to explore how social and gender norms influence VYAs’ SRH in SSA and draw inferences for culturally sensitive, gender-responsive interventions. The review followed the five-step framework developed by Arksey and O’Malley: 1) defining the research question, 2) identifying relevant studies, 3) selecting studies, 4) charting the data, and 5) collating, summarising, and reporting the results. We searched four databases (MEDLINE, CINAHL, Global Health, and Web of Science) for peer-reviewed articles published between January 1, 2000 and December 31, 2024. We identified 24 studies: n=11 (46%) were entirely qualitative, n=8 (33%) were exclusively quantitative, and three other quantitative studies incorporated qualitative components. Two studies used participatory techniques. Studies were from nine countries in SSA. Identified norms included those relating to menstruation, puberty, circumcision, romantic relationships and gender stereotypes. Social norms led to VYAs’ limited SRH knowledge and access, and behaviours and practices that heightened VYAs’ vulnerabilities and poor SRH outcomes. The review also highlighted the interlinkage of various factors that impact VYAs’ SRH, underscoring the need for multifaceted responses. This scoping review underscores the importance of culturally sensitive, gender-responsive interventions in improving the SRH of VYAs in sub-Saharan Africa. It calls for continued research and policy attention to address the complex interplay of social and gender norms, ensuring that VYAs have the necessary resources and support for healthy development. The review provides valuable insights for developing tailored interventions for this important group.

## Introduction

The World Health Organisation (WHO) defines “adolescents” as individuals in the 10–19 years age group [1]. Worldwide, there are ∼1.3 billion adolescents, accounting for 16% of the global population [2]. About half are classified as very young adolescents (VYAs, aged 10–14 years), with around 90% of VYAs residing in low- and middle-income countries [2]. In Africa, the number of young people is projected to double in the next 30 years, rendering (very young) adolescents a particularly important group [3]. As predicted by the World Bank, Africa’s ability to benefit from the projected population growth directly depends on the health and well-being of today’s adolescents and the educational opportunities available to them [3]. It is therefore crucial to invest in the health of VYAs across a wide range of outcomes, which can lead to broader societal gains, including improved productivity and economic gains [4].

In sub-Saharan Africa (SSA), it is particularly critical to focus on VYAs’ sexual and reproductive health (SRH) given adolescents in this region have the worst SRH outcomes of this age group worldwide [5, 6]. For instance, unmarried girls have the highest rates of abortion and abortion-related morbidity and mortality of any region [5, 6]. Rates of curable sexually transmitted infections (STIs), including chlamydia, gonorrhoea, syphilis and trichomoniasis are comparatively higher, and key SRH service indicators such as contraceptive use, antenatal visits or HIV testing are poorer than in any other WHO region [7, 8]. Further, even before the current international HIV funding crisis [9-11], adolescent girls in SSA accounted for 75% of new HIV infections globally [12]. These SRH challenges are due to a range of intersecting factors – behavioural, structural, socioeconomic and sociocultural, including social and gender norms [13, 14].

Social norms are unwritten, informal rules that determine how groups of people ought to behave in certain situations [4, 15, 16]. Gender norms are a subtype of social norms that dictate how males and females ought to behave [4, 15-18]. Social and gender norms are accompanied by sets of “sanctions”, both positive and negative, for norm abiders/adherents and violators alike [16, 19]. Particularly in SSA, social and gender norms are hinged on, and reflect, the predominantly patriarchal character of most societies [16, 18, 20]. Patriarchal structures generally confer power on males to control resources and dominate females, leading to social and gender norms that are favourable to the former and punitive to the latter [21]. There are several other intertwined norms related to sexuality, virginity, fertility, childbearing, and family planning use [20, 22, 23]. These mostly impede healthy SRH behaviours and service-seeking, and contribute to poorer SRH outcomes.

While gender socialisation begins in childhood (<10 years), it intensifies in very young adolescence (10–14 years) and solidifies in later adolescence (≥15 years) [24-26]. However, the plasticity of the VYA adolescent brain offers a prime opportunity to shape self-perception and behaviour, and manipulate social constructs [25]. Intervening in very young adolescence, when attitudes and perceptions are still malleable, provides the opportunity to promote gender-equitable identities and challenge inequitable gender stereotypes before they are solidified and become less amenable to change [4]. Once positive social and gender norms are inculcated, their impact has important consequences for the well-being and SRH of VYAs both now and over their life course [4, 15].

In SSA though, SRH programmes have mostly focused on older adolescents (15–19 years) [27], due to at least three reasons. Firstly, very young adolescence (10–14 years) is generally a time of good health, when this group is not as vulnerable to illness as when they are younger [28]. Secondly, VYAs are less prone to risk-taking than older adolescents [29]. Lastly, there exist widespread misperceptions that VYAs are too young to be sexually active or to need SRH information [30-32], which serve as obstacles to both programme and research development. Consequently, a few initiatives have focused on VYAs, with the Global Early Adolescent Study (GEAS) [33] being an exception.

The GEAS is a multi-country, longitudinal initiative that is exploring VYAs’ perceptions of the gender norms that regulate their behaviour, how they form their own beliefs about gender, and how these beliefs align with social norms in their communities, including in four African countries: Democratic Republic of Congo, Kenya, Malawi and South Africa [34]. An important GEAS observation has been that, as with other population groups, intervention effects do differ by context, and results can be highly contextual, even for settings generally considered “similar” [35]. Indeed, there is increasing recognition that African masculinities are produced in unique and varying contexts of intersections (including ethnicity and sexuality) [36]. Therefore, whilst some core intervention components can be implemented in different settings, there is a need for context-specific adaptations.

To inform such adaptations, we first need to appreciate the breadth and scope of initiatives so far. We therefore conducted a scoping review to explore a body of literature to identify what is known about how social and gender norms influence VYAs’ SRH in SSA to inform culturally sensitive, gender-responsive interventions in Zimbabwe and the region more widely.

## Materials and Methods

### Study design

We conducted an initial search in publicly available registries, including PROSPERO and Open Science Framework (OSF) for planned or ongoing reviews, and did not identify any focusing on this topic. We developed a protocol to guide our review, uploaded it on OSF and subsequently published it [4]. We conducted the review according to the Preferred Reporting Items for Systematic Reviews and Meta-Analyses Extension for Scoping Reviews (PRISMA-ScR) guidelines (S1 File) [37]. We employed the scoping review framework developed by Arksey and O’Malley [38]. The review stages include: 1) defining the research question, 2) identifying relevant studies, 3) selecting studies, 4) charting the data, and 5) collating, summarising and reporting the results [38].

### Stage 1: Defining the research question

We used the Population–Concept–Context (PCC) framework [39] (Table 1) to inform the main review question.

**Table 1:**
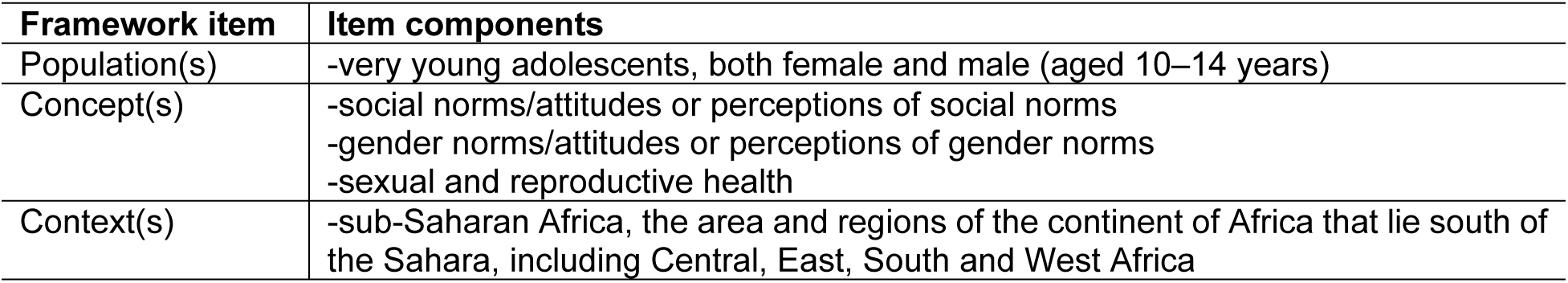
PCC Framework.

The main review question was *"How do social and gender norms influence the SRH of very young adolescents (aged 10–14) in SSA?"* A sub-question was *"To what extent is the influence similar or different for girls and boys?"* These questions enabled us to map the range of relevant literature around these aspects and inform the direction of future research and programming.

### Stage 2: Identifying relevant studies

We developed a search strategy to identify relevant studies written in English from 1 January 2000 to 31 December 2024 (inclusive). We did not have resources to analyse studies written in other languages, and we recognise that this is a potential limitation. We chose 2000 as our baseline year as this is about when research on social and gender norms, VYAs and SRH issues in SSA intensified, especially as part of the Millennium/Sustainable Development Goals’ global health programmes [40]. For example, Sustainable Development Goal (SDG) 5 recognises gender equality as a fundamental human right and a necessary foundation for a peaceful, prosperous and sustainable world [41].

We identified studies relevant to this review by searching electronic databases of published literature in PubMed/MEDLINE, CINAHL, Global Health and Web of Science. We carefully selected these databases for their comprehensiveness in covering the area under research. Our familiarity with these databases also enhanced efficiency in the search process. With the help of a librarian, we first developed the MEDLINE search strategy (S2 File) and later adapted it for the other three databases. Overall, the general search strategy was informed by the PCC framework, as shown in Table 2.

**Table 2:** Search strategy.

| Concept | Search Terms |
| --- | --- |
| Social norms | "Social norms" OR "Cultural pluralism" OR "Social norms attitudes" OR "Social norms perceptions" OR "sociocultural restrictions" OR "protective behaviours" OR "African culture" OR; |
| Gender norms | "Gender" OR "Gender norms" OR "Gender norms attitudes" OR "Gender norms perceptions" OR "Gender roles" OR "Gender attitudes" OR "Gender practices" OR "Gender Transform*" AND; |
| Sexual and reproductive health | "Sexual behaviour" OR "sexual health" OR "Youth sexual behaviour" OR "Attitudes toward sex" OR "Sexuality" OR "Sexual Health -- In Adolescence" OR "Reproductive Health -- In Adolescence" OR "contraceptives" OR "family planning" OR "Protected sex" OR "HIV" OR "STI" OR "sexual transmitted diseases" OR "pregnancy" AND; |
| Very young adolescents | "Youth" OR "young person" OR "minor" OR "10–14 years old" OR "adolescent" OR "teenage" OR "young adolescent*" OR "Very young adolescents" OR "early adolescent" AND; |
| Sub-Saharan Africa | "Angola" OR "Benin" OR "Botswana" OR "Burkina Faso" OR "Burundi" OR "Cameroon" OR "Cape Verde" OR "Central African Republic" OR "CHAD" OR "Comoros" OR "Congo" OR "Congo Democratic Republic" OR "Djibouti" OR "Equatorial Guinea" OR "Eritrea" OR "Ethiopia" OR "Gabon" OR "Gambia" OR "Ghana" OR "Guinea" OR "Guinea-Bissau" OR "Cote d'Ivoire" OR "Ivory Coast" OR "Kenya" OR "Lesotho" OR "Liberia" OR "Madagascar" OR "Malawi" OR "Mali" OR "Mozambique" OR "Namibia" OR "Niger" OR "Nigeria" OR "Sao Tome and Principe" OR "Rwanda" OR "Senegal" OR "Seychelles" OR "Sierra Leone" OR "Somalia" OR "South Africa" OR "South Sudan" OR "Sudan" OR "Swaziland" OR "Tanzania" OR "Togo" OR "Uganda" OR "Zambia" OR "Zimbabwe" OR "Africa, South of the Sahara" OR "sub-Saharan Africa" |

On 19^th^ April 2024, the lead author (FAA) tested a general search strategy by running it using a Boolean search on the Discover platform, combining the search terms from the PCC framework using “AND” and separating different terms using “OR”. She then searched each of the four databases (PubMed/MEDLINE, CINAHL, Global Health and Web of Science) using PCC framework concepts and their synonyms to identify specific terms used in these databases. She then combined the specific terms with free text terms contributed by co-authors. Reviewers then downloaded and imported search results into EndNote 20. After removing duplicates in EndNote 20, they exported the articles to Covidence, a collaborative software. The search was closed on 31^st^ December 2024.

### Stage 3: Study selection

Reviewers removed additional duplicates in Covidence and proceeded with study selection, which consisted of two levels of screening, that is, title and abstract review and full-text review. FAA and a co-reviewer (ON) conducted the two levels of screening using Covidence software. The two reviewers reviewed any discordant full-text articles a second time and further disagreements about study eligibility at the full-text review stage were resolved through discussion with a senior researcher (WM) until full consensus was reached.

### Inclusion and exclusion criteria

Guided by the PCC framework, we included studies meeting all the criteria in each category outlined in Table 3. We included studies that employed quantitative, qualitative or mixed methods. We excluded reviews (scoping, narrative, systematic, meta-analyses, etc.), personal opinion articles, and conceptual or theoretical articles that neither analysed primary nor secondary data. We accounted for all excluded material to appreciate any potential biases or implications of the exclusions to our findings. Table 3 shows the review’s eligibility criteria, including the rationale.

**Table 3:** Studies eligibility criteria.

| Criteria | Inclusion criteria and rationale | Exclusion criteria and rationale |
| --- | --- | --- |
| Population | -Articles reporting on VYAs, both female and male (aged 10–14 years). We focused on VYAs as programmes mostly leave out this group [27]. | -If articles did not report on VYAs at all. |
| Concept | -Articles reporting on social norms/attitudes or perceptions of social norms<br>-Articles reporting on gender norms/attitudes or perceptions of gender norms<br>-Articles reporting on SRH | -Articles not focused on these concepts. |
| Context | -Studies conducted in any country within SSA. | -Studies conducted outside SSA. |
| Study design/type of evidence | -Articles reporting findings from primary research.<br>-Articles published in English.<br>-Articles published between 1st January 2000 and 31<sup>st</sup> December 2024. | -Reviews (e.g., systematic or scoping reviews)<br>-Non-English articles as we did not have the resources to analyse articles published in other languages.<br>-Articles published before 1st January 2000. |

### Stage 4: Data charting

Data charting is the process of synthesising and interpreting data through sorting, charting and organising information based on key themes and issues [38]. In line with Levac et al.’s [42] recommendations, we developed a standard Excel data charting form and populated it with key study characteristics (e.g., author(s), publication year, country/geographical region, study design, study population), and key issues from each study in relation to review objectives. This was an iterative process, involving back-and-forth data extraction and subsequent updating of the data charting form.

We initiated the data charting process with a pilot phase, where two independent reviewers (FAA and ON) charted data from a random sample of 5-10 of the final selected studies. This piloting process determined whether the independent chartings of both reviewers aligned with the review objectives and allowed for any necessary changes to the data charting form. Any disagreements were discussed with a more senior researcher (WM), and the data charting form was revised accordingly.

### Stage 5: Collating, summarising and reporting the results

We used descriptive statistics to present the characteristics of the included studies. Specifically, we presented the number of studies meeting study criteria (e.g., geographical location, study period, study design). Following best practices in thematic analysis, which include being purposefully iterative and reflexive [43], we then thematically examined the main conclusions drawn from the included studies. Before logically developing and applying preliminary codes and themes to the data retrieved from the chosen studies, FAA and ON first carefully reviewed and comprehended the extracted data. We then developed an initial codebook.

To continuously improve the thematic analysis process, we discussed and shared notes throughout. For instance, we were able to create new codes and themes or combine existing ones when there were overlaps. To better comprehend our findings and pinpoint pertinent gaps in relation to our review questions, we finally merged all of our studies and mapped out the similarities and differences in our data [44]. This final analysis was developed and refined with input from all co-authors.

### Ethics and dissemination

Ethical approval was not required as this was a literature review.

## Results

### Search outcomes

The search yielded 1,912 potentially relevant records from the four databases. Covidence detected 795 duplicate records, and two additional duplicates were removed manually. There remained 1,115 records for title and abstract screening. Title and abstract screening resulted in the exclusion of an additional 1,084 articles, which did not fit the PCC framework components.

There remained 31 articles for full-text review. After cross-validation and full-text screening, we included 24 articles in the review. There was some considerable discussion on whether or not to include one study which investigated VYAs’ SRH experiences from the perspective of emerging adults (18–25 years old) [45]. In the end, the study was included as it met all the inclusion criteria, despite not directly involving VYA participants. Fig 1 presents a summary of the selection process.

**Fig 1:**
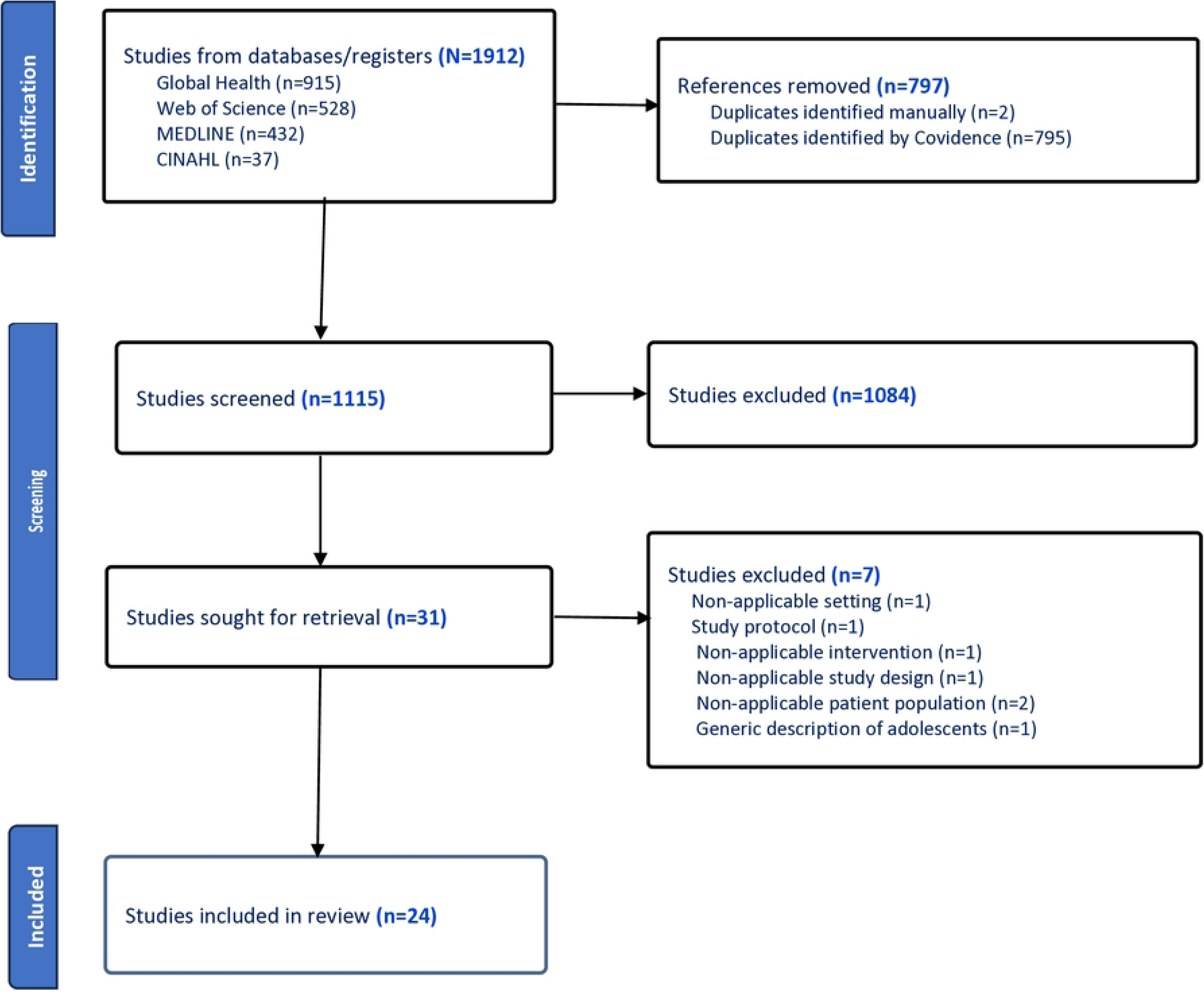
PRISMA flowchart of study selection process.

### Study descriptions

The 24 studies included for final review were conducted in nine SSA countries, and distributed across East, South and West Africa, with about two-thirds (n=16, 67%) from East Africa (Table 4). Of the 24 studies, n=7 (29%) were conducted in Uganda, n=6 (25%) in Kenya, with one study [46] in both Kenya and Nigeria. Malawi and the Democratic Republic of the Congo had three studies each. A multi-country study [47] was conducted in Burkina Faso, Ghana, Malawi and Uganda. South Africa and Ethiopia had two studies each, with one of the studies conducted in Ethiopia [48] focusing on Somali refugees. Whilst the review period was 2000-2024, the earliest article was published in 2007. All other papers were published in the last 15 years (i.e., 2009-2024) (Table 4).

**Table 4:** Study characteristics.

|  | Authors | Country | Study design | Study population | Sample size | Key findings on social & gender norms and SRH |
| --- | --- | --- | --- | --- | --- | --- |
| 1 | Baird, S. et al., 2022 | Ethiopia | Mixed methods (Quan & Qual) | Boys & Girls | 2,879 | Menstrual stigma and shame; menstruation associated with a curse.<br>Boys view menstruation as “wrong”, teasing girls during periods.<br>Sex education perceived as encouraging promiscuity. |
| 2 | Bankole, A. et al., 2007 | Burkina Faso, Ghana, Malawi, Uganda | Quantitative | Boys & Girls | 8,837 | VYAs already sexually active and many believe their close friends are too.<br>VYAs had high awareness levels, but little in-depth knowledge of pregnancy and HIV prevention.<br>Multiple information sources used and preferred. |
| 3 | Bello, B.M. et al., 2017 | Kenya, Nigeria | Qualitative | Boys & Girls | 66 | VYAs' reactions to bodily changes vary from anxiety to pride.<br>VYAs tend to desire greater privacy, trying to hide their developing bodies from others.<br>Girls emphasise breast development as the marker of puberty, while boys emphasise voice changes.<br>Among some ethnic groups in Nairobi male circumcision viewed as the hallmark of adolescence. |
| 4 | Bunoti, S.N. et al., 2022 | Uganda | Qualitative | Boys & Girls | 152 | Rural girls more aware of their body changes than urban ones.<br>Urban boys more knowledgeable of pubertal body changes than rural ones.<br>Girls use herbs as pain killers to reduce menstrual pain.<br>Poor pubertal hygiene practices such as shaving due to limited knowledge and resources.<br>Poor parent-child communication related to sex and puberty. |
| 5 | Chimwaza-Manda, W. et al., 2021 | Malawi | Qualitative | Girls | 23 | Due to gender norms, VYA girls have fewer opportunities than boys to socialise and engage in leisure and income-generating activities.<br>Gender differences where household chores done by girls and boys left to play. |
| 6 | Chimwaza-Manda, W. et al., 2023 | Malawi | Qualitative | Girls | 43 | Parents and adult relatives often advise against engaging in sexual relationships to prevent pregnancy, preventing some VYA girls from discussing SRH issues.<br>VYA rely on peers' incorrect information. |
| 7 | Chimwaza-Manda, W. et al., 2024 | Malawi | Qualitative | Girls | 43 | VYA girls' social dynamics are influenced by their trust and confidence in SRH information sources.<br>Parents/caregivers only start to talk to VYA girls about SRH issues (e.g., prevention of pregnancy) when they have already started menstruating. |
| 8 | Cislaghi, B. et al., 2021 | DRC | Quantitative | Boys & Girls | 2,163 | Changes in Sexual Double Standard (SDS) scores are linked to puberty and pubertal onset, with girls facing increased pressure to maintain their sexual purity.<br>Boys reported greater freedom of movement, spent more time with peers, and socialised in mixed-sex groups. |
| 9 | Gayles, J. et al., 2023 | DRC | Mixed methods (Quan & Qual) | Boys & Girls | 2,519 | Discriminatory attitudes towards gender nonconforming adolescents.<br>Unequal gender norms, such as household chore sharing. |
| 10 | Kagesten, A.E. et al., 2018 | Kenya | Quantitative | Boys & Girls | 365 | Parents disapprove of sexual relationships in VYAs for both boys and girls.<br>Masculine norms become stereotypical in early adolescence. |
| 11 | Kemigisha, E. et al., 2018a | Uganda | Quantitative | Boys & Girls | 1,096 | • SRH knowledge among VYAs is poor.<br>• There are significant gaps in information about early sexual debut and risky practices. |
| 12 | Kemigisha, E. et al., 2018b | Uganda | Quantitative | Boys & Girls | 1,096 | • VYAs socialised at a young age to have inequitable norms<br>• Boys perceived as cleverer, and girls expected to stay home.<br>• Girls had higher gender equitable norm scores. |
| 13 | Kemigisha, E. et al., 2019 | Uganda | Mixed methods (CRT & Qual) | Boys & Girls | 1,116 | • Sexual wellbeing and attitudes based on self-esteem and body image and gender equitable norms. |
| 14 | Maina, B. et al., 2020a | Kenya | Quantitative | Boys | 426 | • Young people begin being romantically involved at an early age.<br>• VYAs understand social norms and power dynamics in heteronormative romantic relationships, but they do not apply these beliefs to their sexual activities.<br>• Boys' endorsement of heteronormativity in romantic relationships was low, suggesting they disagreed with traditional gender judgments<br>◦ However, their endorsement of male dominance in romantic relationships had a significant inverse association with sexual experiences. |
| 15 | Maina, B. et al., 2020b | Kenya | Quantitative | Girls | 606 | • Girls who had experienced violence were more likely to be sexually experienced.<br>• Self-reported depressive symptoms were associated with sexual experiences among VYA girls, leading to poor impulse control, impaired decision-making and increased risk of engaging in risky sexual behaviours. |
| 16 | Maina, B. et al., 2020c | Kenya | Qualitative | Boys & Girls | 30 | • Parents discourage romantic relationships among VYAs, viewing them as an obstacle to educational achievement.<br>• Lack of communication about sexual relationships is primarily due to the perception that children are too young to engage in romantic relationships.<br>• Fear-based communication, including threats, is employed to prevent romantic relationships.<br>• Discussing sex-related matters with adolescents may be interpreted as approval or tolerance of promiscuity. |
| 17 | Maina, B. et al., 2022 | Kenya | Qualitative | Boys | 27 | • Boys observe older men's behaviours and roles, receive messages on masculine expectations from parents and teachers.<br>• Contextual masculinity is associated with circumcision, and Muslim boys perceive masculinity based on religious teachings.<br>• Media and internet messages portray men as expected to support and take family financial responsibilities. |
| 18 | Mda, P. et al., 2013 | South Africa | Qualitative | Girls | 25 | • Parents fear discussing contraceptives and romantic relationships with VYA girls.<br>• Older girls responsible for teaching young girls once they reach puberty.<br>• Adolescent pregnancy rates are linked to ignorance about contraception, coercion, rape and low modern contraceptives access. |
| 19 | Ninsiima, A.B. et al., 2018 | Uganda | Qualitative | Boys & Girls | 56 | • Long-term gender ideals include girls' values for motherhood, marriage and caregiving.<br>• Social norms require parents and teachers to control girls more than boys.<br>• Girls are expected to control their own sexuality and take responsibility for boys' sexualities; prevention of pregnancy is perceived as a girl's responsibility. |
|  |  |  |  |  |  | Boys are generally independent and have unlimited mobility, while girls are socially expected to be submissive and have restricted movements.<br>Sexuality education mainly focuses on biological aspects like hygiene and control of girls' sexuality.<br>Gender norms form early in life and create unequal power relations that constrain adolescents from exercising agency. |
| 20 | Nyakato, V.N. et al., 2021 | Uganda | Participatory | Boys & Girls | 20 | When a girl gets pregnant, it brings shame to the family and more particularly, the mother and other girls.<br>Traditional cultural norms emphasise gender differences, with boys working on farms, plantations, and gardens, while girls maintain the household and care for younger siblings. |
| 21 | Ortiz-Echevarria, L. et al., 2017 | Somali refugees in Ethiopia | Qualitative | Boys & Girls | 62 | Inequitable relations between boys and girls, and harmful traditional practices speak to the lack of self-efficacy and decision-making that VYAs and specifically girls, experience.<br>Girls faced an additional risk of child marriage and early pregnancy, with displacement significantly limiting their ability to access education and achieve future aspirations. |
| 22 | Selikow, T.A. et al., 2009 | South Africa | Qualitative | Boys & Girls | 8 FGDs | Limited access to adults, knowledge and hence the adolescents rely on peers for information.<br>Adolescents often feel a sense of belonging in groups, which can be exploited to promote negative sexual norms.<br>Peer pressure among both boys and girls undermines healthy social norms and HIV prevention messages to abstain, be faithful, use a condom and delay sexual debut. |
| 23 | Tchuisseu et al., 2023 | Uganda | Mixed methods (participatory & FGDs) | Boys & Girls | Participatory activities plus n=20 | Girls are expected to avoid pregnancies.<br>Girls receive more warnings and stay at home, while boys are not always expected to stay at home.<br>Limited economic resources, sex education and gender expectations can lead to negative SRH outcomes.<br>School and peers provide sex education, but girls are more likely to experience rape and transactional sex. |
| 24 | Zimmerman, L.A. et al., 2021 | DRC | Quantitative | Boys & Girls | 2,610 | An increase in sexual double standards among adolescents who have not yet reached puberty.<br>While no significant sex differences exist in decision-making power, boys have greater voice and representation. |

### Study characteristics

Of the 24 studies, n=11 (46%) were exclusively qualitative [46, 48-57] and n=8 (33%) were entirely quantitative [47, 58-64]. Two other quantitative studies [65, 66] and the only cluster-randomised trial [67] incorporated qualitative components. One study [68] exclusively used participatory techniques while the other [45] used these alongside focus group discussions. Overall, n=17 (71%) studies explored social norms and SRH among both VYA boys and girls, n=5 (21%) among only girls and n=2 (8%) among only boys (Table 4). Key reported issues are summarised in Table 4 and described in detail below.

### Key reported issues

#### Beliefs, practices and VYAs’ SRH

The review identified several norms linked to VYA’s SRH, including menstrual-related norms and associated stigma, shame and limited mobility [65]. Adult females only discussed menstruation with VYA after it had commenced. Additionally, discussions rarely focused on SRH-related aspects but rather, on the need to maintain hygiene and to use traditional remedies to manage associated pain (e.g., herbs) [49]. In some settings, male circumcision was associated with masculinity when uncircumcised boys were both looked down upon and ridiculed, with some eventually getting circumcised to both gain social acceptance and avoid stigmatisation [56]. A study conducted in South Africa illustrated how hegemonic traditional masculine norms sometimes led to negative sexual norms when non-sexually experienced VYA boys were labelled *“umqwayito”* (dried fruit or meat) – derogatory terms which forced some to engage in (sometimes risky) sexual activity to conform to group norms [55].

#### Parental communication and gatekeeping

Overall, parents, caregivers and other adults believed that VYAs were too young to engage in romantic relationships and therefore avoided discussing SRH issues with them [51, 63]. Consequently, VYAs had to rely on their peers and the media for SRH information [55], but often found it challenging to determine the information’s accuracy [56, 68]. They were therefore prone to myths and misconceptions. For example, in one study, VYAs reported fear of using contraceptive pills due to perceptions that they have long-term effects, including infertility [53].

Additionally, parents perceived any romantic interactions during very young adolescence as a hindrance to academic achievement [63]. Others regarded such relationships as disgraceful to the family. Consequently, parents used threats to deter such relationships, which in turn, resulted in VYAs feeling scared to either discuss or disclose their feelings and experiences, leading to a breakdown in healthy communication [57].

There was evidence of differential treatment between VYA boys and girls during the pubertal stage when carers became more concerned about the girls’ sexuality. This often led to heightened expectations for VYA girls to behave in a respectable manner and maintain their sexual purity to prevent bringing shame upon their families [58]. This was compounded by the general fear that girls who fall pregnant may not be accepted by the perpetrator’s family [68]. Of note, girls were expected to simultaneously exert social control over their own and boys’ sexuality due to the perception that pregnancy is primarily a girl’s responsibility [68].

#### Gender norms, beliefs and VYAs’ SRH

Due to concerns around VYA girls’ heightened vulnerability, parents and other stakeholders (e.g., teachers) instituted stricter measures on VYA girls compared to boys [54]. For example, study participants mentioned that VYA boys enjoyed free movement and could have multiple, concurrent partners while girls’ sexuality was restricted [54, 59]. Girls who expressed sexual agency were perceived as loose or abnormal [54]. Boys maintained that in heterosexual romantic relationships, they were supposed to be dominant, with girls expected to be passive [59, 64].

Two studies [58, 64] explored a specific gender norm: the Sexual Double Standard (SDS), a measure of different normative expectations for romantic activities, rewarding boys but devaluating girls for engaging in the same behaviours [27]. The studies found a high SDS prevalence among VYAs, which increased over time, and was influenced by pubertal onset, family interactions, peer interactions and media exposure [58, 64]. In one of the studies, VYA boys agreed that boys and girls should face different judgements for the same sexual behaviour [58].

#### Contextual factors

From the review, it was evident that contextual factors have a bearing on VYAs’ SRH outcomes. For example, living in urban informal settlements led to comparatively higher common mental disorders, including anxiety and depression as these settings exhibit significant social, environmental and physical risks [59]. Self-reported depressive symptoms were associated with sexual experiences among VYA girls, leading to poor impulse control, impaired decision-making and increased risk of engaging in risky sexual behaviours [62]. Studies also demonstrated that factors at various level impact VYAs SRH. For example, household dynamics, including parental loss may result in early sexual debut [50]. At the community level, sociocultural factors including ethnicity, cultural practices, gender roles, cultural beliefs and safety impact VYAs’ SRH [48, 64].

## Discussion

This review explored how social and gender norms influence the sexual and reproductive health (SRH) of very young adolescents (VYAs, 10–14 years) in sub-Saharan Africa (SSA). All but one of the studies included in the review were conducted in the last 15 years, reflecting an increasing but comparatively nascent interest in this age group.

Identified norms include those relating to menstruation, puberty, circumcision, romantic relationships and gender stereotypes. Overall, norms restrict VYAs girls’ access to SRH information and services. A recurring theme is that parents, teachers and services providers continue to believe that VYAs are too young to be sexually active or to need SRH information [30-32]. VYA therefore end up relying on the media and their peers for SRH information [55]. Peers do not often have adequate or correct information; this is also true for older adolescents. A study among girls aged 15–19 years in Uganda [69] found that even these older adolescents have challenges accessing SRH information. This highlights a common gap among adolescents regardless of their age, and the need to provide this group with adequate, correct SRH information. One of the reasons for the gap could be because VYAs are often seen as children and older adolescents as adults [33]. There is need to recognise adolescents as a distinct group, with own specific needs and issues, and provide appropriate SRH information.

The review identified parental challenges around discussing SRH issues with VYA, even if they wanted to do so. Without adequate knowledge and skills, the communication predominantly involves conveying warnings, issuing threats and resorting to physical discipline [57]. However, recent research on the impact of parenting on adolescent sexual risk-taking has demonstrated that supportive parent-child relationships can reduce the risk of adolescents engaging in unprotected sexual activities [70]. Interventions targeting VYAs’ SRH should therefore include a component focusing on parent-child relationships. In fact, in keeping with the socioecological model, which shows how individuals are embedded within larger social systems [71], interventions need to focus on the wider support system. It will also be important to identify and explore the community resources and structures available to ensure sustainability and scalability.

Among VYA boys, both peer pressure and the need to conform to group norms are linked to SRH behaviours and practices. Of concern, however, is the influence of hegemonic traditional masculine norms which sometimes result in VYA boys engaging in aggressive sexual activity [55]. Conversely, in some reviewed studies, VYA girls reported sexual coercion and rape [45, 53].

Adolescents in general and YYA, in particular assign greater weight than adults to social outcomes such as peer acceptance [72]. Previous research has characterised the development of vulnerability to peer influence during adolescence as a pattern that resembles an inverted U-shaped curve, showing an initial rise during early adolescence, reaching its highest point around 14 years of age, and thereafter decreasing [73]. VYAs therefore exhibit lower levels of resistance to peer pressure compared to both children and adults [73]. Interventions focusing on VYA should therefore include life skills to deal with peer pressure. Gender transformative approaches, that work with groups of males and females to promote critical reflection on harmful gender norms and unequal power dynamics, and build relationship skills such as communication and conflict resolution [18], can empower VYAs to challenge harmful norms.

One the whole, however, VYAs’ SRH is influenced by a complex interplay of social, cultural, and economic factors. For example, among adolescents in impoverished areas (e.g., slums), VYAs’ SRH is influenced by numerous intersectional factors such as crime, prostitution, violence and substance use [16, 63]. These factors may shape for example, VYAs’ masculinities but importantly, affect their mental well-being. Reviewed studies suggested a link between poor mental health and SRH outcomes. There is a recognised bidirectional relationship between the two in that poor mental health leads to poor SRH outcomes and vice versa [74]. Further, depression, HIV/AIDS and self-harm are three of the top five global causes of disability-adjusted life years lost for adolescents [75]. Interventions need to recognise these linkages and avoid the tendency to focus on just one aspect and not the other. Above all, given the various intersecting factors, interventions need to be careful not to over-emphasise individual determinants of health, overlooking other factors that shape life chances, health risks, and vulnerabilities [76].

### Strengths and limitations

This scoping review allowed us to broadly examine how social norms influence the SRH of VYAs in SSA. The strength of the review is in the application of a recognised, thorough and transparent approach [38] to review the literature and report our results. We also employed a two-stage, double screening process to minimise bias. However, given that a significant part of SSA is Francophone or Lusophone, and programmes in these regions are disproportionately funded [4], this is likely to skew study results. This may have been compounded by the inclusion of only articles published in English (as we did not have the resources to analyse studies in other languages). Finally, most of the issues reported here relate to heterosexual VYAs. Future research should explore similar issues among VYAs from other settings and sub populations (e.g., gender non-conforming VYAs).

## Conclusions

This scoping review underscores the importance of culturally sensitive, gender-responsive interventions in improving the SRH of VYAs in sub-Saharan Africa. It calls for continued research and policy attention to address the complex interplay of social and gender norms, ensuring that VYA have the resources and support they need for healthy development. The review provides valuable insights for the development of tailored interventions for this important group.

## Data Availability

All data underlying the findings are included in the manuscript.

## Acknowledgments

We would like to acknowledge the support that was offered by Alison Derbyshire and Maria Angelica Rweyemamu in search terms development and online library system manoeuvring.

## Supporting information

S1_Table: Preferred Reporting Items for Systematic reviews and Meta-Analyses extension for Scoping Reviews (PRISMA-ScR) Checklist

S2_Fig: MEDLINE search

